# A Dual-Species Plant-derived Extracellular Vesicle and Growth Factors Formulation for Hair and Scalp Care: A Randomized, Placebo-controlled Study

**DOI:** 10.64898/2026.09.07.26362401

**Authors:** Tsong-Min Chang, Chung-Chin Wu, Huey-Chun Huang, Ji-Ying Lu, Yu-San Chen, Pei-Lun Kao, Wei-Hsuan Tang, Wang-Ju Hsieh, Luke Tzu-Chi Liu, Ivona Percec, Charles Chen, Tsun-Yung Kuo

## Abstract

Scalp dysregulation and hair shedding are common cosmetic concerns, driving an increasing demand for non-pharmaceutical interventions, including plant-derived extracellular vesicles (EV) and growth factors. This randomized controlled trial evaluated the efficacy of topical formulations in 60 healthy adults over a 56-day period, monitoring scalp and hair parameters at Days 14, 28, 42, and 56 relative to baseline (Day 0). Group A: placebo; Group B: base (placebo containing 0.1% caffeine and panthenol); Group C: base + recombinant Fc-fusion long-acting insulin-like growth factor-1 (rIGF-1) and fibroblast growth factor-7 (rFGF-7); Group D: base + *Centella asiatica* (*C. asiatica*) + ginger EVs; and Group E: base + rIGF-1 and rFGF-7 (each at twice the concentration in Group C), and *C. asiatica* + ginger EVs. Group E demonstrated rapid onset and significant superiority over the control and other treatment groups across all parameters. In scalp regulation, Group E achieved a highly significant, time-dependent reduction in excess sebum, reaching a 67% decrease by Day 56. Hair growth kinetics were significantly accelerated in Group E, with a mean cumulative length increase approaching 3.9 cm by Day 56, driven by a surge in mean weekly growth velocity to 1.4 cm/week. Group E also demonstrated the highest improvements in hair thickness (19 µm) and hair density (17% increase) by Day 56. Finally, hair shedding was reduced by almost half by Day 56 in Group E. These results demonstrate that the formulation delivers a clinically superior non-pharmaceutical intervention for advanced hair and scalp revitalization.

## Introduction

Hair plays a central role not only in scalp protection and thermoregulation but also in personal appearance, self-confidence, and social interactions_[1,2]_. Because of this, hair thinning and hair loss often cause substantial psychological distress and reduced quality of life_[3,4]_. Although numerous cosmetic and pharmaceutical approaches have been developed to address hair loss, maintaining a healthy scalp environment that continuously supports hair follicle function remains a major challenge_[5-8]_.

Hair growth depends on the ability of hair follicles to repeatedly cycle through the anagen, catagen, and telogen phases_[1,2]_. During anagen, active proliferation and differentiation of hair matrix keratinocytes drive hair shaft elongation, thickness, and density _[1,2]_. This process is tightly regulated by complex interactions among dermal papilla cells, follicular epithelial cells, growth factors, immune mediators, and the surrounding scalp microenvironment_[5-8]_. Consequently, modern scalp-care strategies increasingly focus on supporting the broader follicular ecosystem rather than targeting a single biological pathway_[5-8]_.

Among the numerous signaling molecules involved in hair biology, insulin-like growth factor-1 (IGF-1) and fibroblast growth factor-7 (FGF-7, also known as keratinocyte growth factor) are recognized as key regulators of follicular activity_[9-14]_. IGF-1 promotes follicular cell survival, inhibits apoptosis, and prolongs the anagen phase_[9-11]_, whereas FGF-7 stimulates keratinocyte proliferation and contributes to hair follicle development and regeneration_[12-14]_. Together, these growth factors provide complementary biological support for maintaining active hair growth_[9-14]_.

More recently, extracellular vesicles (EVs) have emerged as a promising class of bioactive entities capable of mediating intercellular communication_[15-18]_. Plant-derived EVs, in particular, have attracted increasing attention because they naturally carry proteins, lipids, nucleic acids, and other signaling molecules that can influence tissue repair, inflammation, and cellular homeostasis_[15-18]_. Their excellent biocompatibility and natural origin make them especially attractive candidates for cosmetic and scalp-care applications_[15-18]_.

Among various plant-derived EVs, ginger (*Zingiber officinale*)-derived extracellular vesicles (GDEVs) have recently gained attention for their potential role in hair regeneration_[15,16]_. Emerging evidence suggests that GDEVs can promote recovery of alopecic lesions and stimulate hair growth in experimental animal models, indicating that ginger may provide more than traditional antioxidant and anti-inflammatory benefits_[15]_. Instead, its extracellular vesicles may actively participate in creating a follicular microenvironment favorable for hair growth_[15,16]_. These findings raise the intriguing possibility that GDEVs could complement conventional growth factor-based strategies by introducing additional biological signals that support follicular recovery and tissue renewal_[15,16]_.

*Centella asiatica*-derived EVs represent another promising plant-derived vesicle system_[17,18]_. *C. asiatica* has long been valued for its wound-healing and anti-inflammatory properties, while recent studies have demonstrated that both its extracts and extracellular vesicles can enhance dermal papilla cell function and promote tissue repair_[17,18]_. In our previous exploratory randomized human study, a scalp essence containing recombinant IGF-1, recombinant FGF-7, and *C. asiatica* EVs significantly improved hair length, diameter, density, scalp oiliness, and hair shedding after 56 days of use_[19]_. These findings provided clinical evidence supporting the concept that combining growth factors with plant-derived EVs may benefit scalp and hair health_[19]_.

Building upon these encouraging results, we sought to further strengthen this multi-target strategy by incorporating ginger-derived EVs into the formulation_[15,16,19]_. In addition, D-panthenol (vitamin B5), a well-established moisturizing and follicle-supporting ingredient, and caffeine, a commonly used hair growth-promoting active known to stimulate follicular proliferation and counteract androgen-related inhibitory effects, were included to provide additional support for scalp barrier function and follicular activity_[20-27]_.

Therefore, the present study evaluated a novel scalp-care formulation containing IGF-1, FGF-7, GDEVs, *C. asiatica* EVs, vitamin B5, and caffeine_[11,14–16,19-27]_. We hypothesized that this next-generation multi-component formulation would establish a superior scalp microenvironment by concurrently supporting follicular cell survival, epithelial proliferation, tissue homeostasis, barrier function, and key pro-growth biological signaling pathways-ultimately resulting in superior clinical improvements across hair and scalp parameters in human subjects.

## Materials and Methods

### Participants and Study Design

This study was a prospective, randomized, placebo-controlled, double-blinded clinical trial conducted to evaluate the effects of a topical scalp revitalizing essence on hair growth and hair loss. Participants were healthy adults of any gender aged 18 to 60 years (inclusive).

Inclusion criteria include healthy adults defined as individuals without chronic diseases, significant illnesses (including cancer, post-stroke disorders, paralysis, acute myocardial infarction, coronary artery bypass surgery, end-stage renal disease, and major organ transplant or hematopoietic stem cell transplant), or active allergic conditions, who are not currently taking medication or using specialized scalp-care products.

Exclusion criteria were defined as: pregnant or breastfeeding individuals; males or females with chronic diseases, major illnesses, or allergic constitutions, current users of medicated scalp products, students or employees directly affiliated with the principal investigator, individuals with scalp abrasions or wounds, those who have participated in other cosmetic trials; or those who had undergone aesthetic scalp procedures within the past three months. Participants were randomly assigned to five parallel groups (n = 12 per group) as detailed below.

Participants were asked to apply approximately 1 mL of the intervention assigned once daily in the evening after shampooing and hair cleaning. The product was applied over the entire scalp using a cotton pad and gently tapped into the scalp with fingertips to promote absorption. Participants were asked not to change their daily hair care routine other than the application of the test product. Usage was self-recorded in a provided diary. The duration of intervention was 8 weeks (56 days), with measurements taken at baseline (Day 0) and on Days 14, 28, 42, and 56.

*C. asiatica* and *Z. officinale* extracellular vesicles were isolated using a previously described centrifugation and filtration method [18]. rIGF-1 (INCI ID: 40916) and rFGF-7 (INCI ID: 40201) are recombinant proteins modified for long-acting by fusing human IGF-1 or human FGF-7 to a human IgG1 Fc region fragment with a flexible linker described previously[19]. The composition of the formulations is as in the following Table 1:

**Table 1.** Composition of the formulations. Plus (+) or minus (−) sign indicates the presence or absence of the component in the formulation.

|  | **Contained in Formulation/Concentration** |  |  |  |  |
|---|---|---|---|---|---|
| **Component** | **A** | **B** | **C** | **D** | **E** |
| *C. asiatica* EV | **−** | **−** | **−** | 10<sup>9</sup> particles/mL | 10<sup>9</sup> particles/mL |
| *Z. officinale* (ginger) EV | **−** | **−** | **−** | 10<sup>9</sup> particles/mL | 10<sup>9</sup> particles/mL |
| rIGF-1 | **−** | **−** | 250 ng/mL | **−** | 500 ng/mL |
| rFGF-7 | **−** | **−** | 150 ng/mL | **−** | 300 ng/mL |
| Caffeine | **−** | **+** | **+** | **+** | **+** |
| Panthenol (Vitamin B5) | **−** | **+** | **+** | **+** | **+** |
| Glycerin | **+** | **+** | **+** | **+** | **+** |
| Capryloyl Glycine | **+** | **+** | **+** | **+** | **+** |
| Xylitylglucoside | **+** | **+** | **+** | **+** | **+** |
| Hexylene Glycol | **+** | **+** | **+** | **+** | **+** |
| Propanediol | **+** | **+** | **+** | **+** | **+** |
| Caprylhydroxamic acid | **+** | **+** | **+** | **+** | **+** |
| 1,2-Hexanediol | **+** | **+** | **+** | **+** | **+** |
| Xanthan Gum | **+** | **+** | **+** | **+** | **+** |
| Menthol | **+** | **+** | **+** | **+** | **+** |
| Menthyl Lactate | **+** | **+** | **+** | **+** | **+** |
| PPG-10 Methyl Glucose Ether | **+** | **+** | **+** | **+** | **+** |
| Aqua | **+** | **+** | **+** | **+** | **+** |

### Randomization and Blinding

Participants were randomly assigned to five parallel groups (n = 12 per group) in 1:1:1:1:1 ratio using block randomization with a fixed block size of five, without stratification. The random allocation sequence was generated by the principal investigator and implemented using sequentially numbered, opaque, sealed envelopes prepared by an independent staff member who was not involved in participant recruitment, assessment, or intervention delivery. The envelopes, identical in appearance and sealed with tamper-proof tape, were stored in a locked cabinet accessible only to the designated unblinded coordinator. Allocation was revealed only after confirmation of eligibility and receipt of written informed consent.

A double-blind design was employed for this study. Participants, care providers administering the interventions, outcome assessors, and data analysts remained blinded to group allocation until the database lock, except in cases requiring emergency unblinding for safety reasons. Blinding was maintained by preparing all intervention and control products in identical containers with matched appearance, labeling, texture, viscosity, and scent. Each product was labeled only with a participant identification number, without an indication of group assignment.

### Assessment of Outcomes

Measurements were performed onsite at Hungkuang University (Taichung City, Taiwan) using two non-invasive diagnostic systems at standardized scalp sites (left, right, and vertex):

Scalp sebum levels were quantified using C+K Multi Probe Adaptor MPA580 system with Sebumeter® SM815 probe (Courage-Khazaka Electronic GmbH, Koln, Germany) via grease-spot photometry.

Hair length, thickness, and density were assessed with the ScalpX Intelligent Scalp Diagnostic System (VAST Technologies Inc., Taipei City, Taiwan) with a 5-megapixel handheld USB digital microscope (DMC1213_USB 200X, Coresync Technology Corp., New Taipei City, Taiwan), and hair loss was assessed by previously described 60-stroke combing test[19].

For sebum content, hair density, and hair loss, values were normalized as percentage improvement over baseline. For hair length and hair thickness, absolute changes (in mm or _μ_m) from baseline were used and for hair length, the mean weekly change in hair length was calculated by subtracting the mean hair length between two timepoints.

Results for each participant were reported and calculated as the average of measurements from the left, right, and vertex regions of each participant.

### Statistical Analysis

Statistical analyses were performed using the built-in statistical package of GraphPad Prism 6.01 (Boston, MA, USA). Results were presented in mean values with 95% confidence intervals. For comparing demographic baseline values, the _χ_2 test was used for gender and the one-way ANOVA was used for continuous variables. To compare within-group changes over time, repeated-measures ANOVA with the Greenhouse-Geisser correction for sphericity, followed by Tukey’s test for multiple comparisons, was used. For between-group comparisons at each time point, one-way ANOVA followed by Tukey’s test was used for multiple comparisons. A p-value < 0.05 was considered statistically significant. All tests were two-tailed. Sample size was determined based on exploratory pilot trial conventions for cosmetic formulations rather than formal power calculations.

## Results

### Participants

Between November 1, 2025, and May 31, 2026, 60 healthy adult participants were enrolled and randomly assigned to one of the five groups with 12 participants per group. There was no attrition of the participants and all completed the study without major protocol violations or deviations. Figure 1 shows the CONSORT flow diagram summarizing the participant flow of the study. Table 2 summarizes the baseline demographic characteristics (presented in raw values).

**Figure 1.**
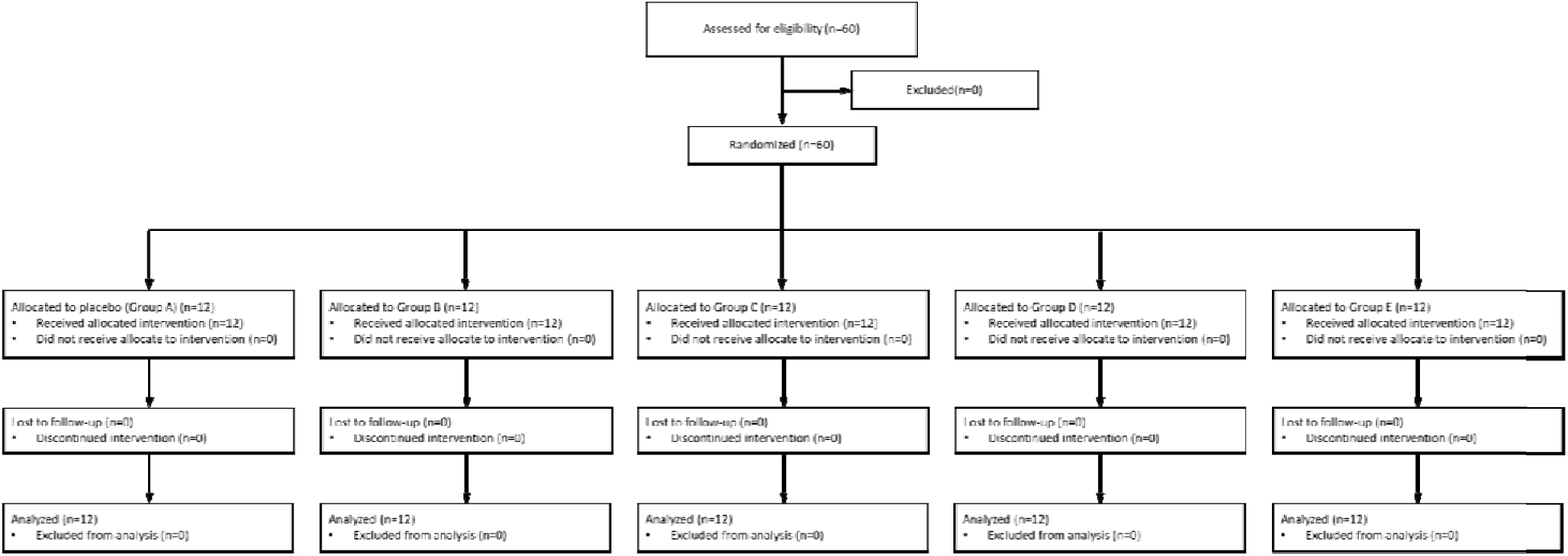
CONSORT flowchart of the study.

**Table 2.** Demographic characteristics and baseline values of the participants.

| **Allocated Group** | **A** | **B** | **C** | **D** | **E** | ***p*-Value \*** |
|---|---|---|---|---|---|---|
| ***n*** | 12 | 12 | 12 | 12 | 12 |  |
| **Gender** |  |  |  |  |  |  |
| **Female** | 10 (83.3%) | 10 (83.3%) | 10 (83.3%) | 11 (91.7%) | 11 (91.7%) | 0.9295 |
| **Male** | 2 (16.7%) | 2 (16.7%) | 2 (16.7%) | 1 (8.3%) | 1 (8.3%) |  |
| **Ethnicity: Asian** | 12 (100%) | 12 (100%) | 12 (100%) | 12 (100%) | 12 (100%) | - |
| **Baseline values (mean ± SD)** |  |  |  |  |  |  |
| Age (years) | 33.3 ± 11.7 | 37.4 ± 11.3 | 34.7 ± 9.3 | 36.0 ± 8.9 | 38.4 ± 12.5 | 0.7880 |
| Sebum content (a.u.) | 90.1 (65.7) | 106.7 (81.0) | 102.0 (68.5) | 140.6 (84.3) | 109.7 (68.1) | 0.5465 |
| Hair length (cm) | 39.2 (13.3) | 37.0 (16.9) | 33.1 (9.9) | 36.3 (13.4) | 43.7 (15.2) | 0.4398 |
| Hair thickness (µm) | 96.6 (8.2) | 91.9 (9.9) | 92.2 (10.5) | 84.8 (10.2) | 85.1 (9.0) | 0.0158 |
| Hair density (no. of hairs/cm<sup>2</sup>) | 186.4 (24.2) | 172.1 (21.7) | 163.2 (18.7) | 162.8 (16.6) | 168.6 (15.0) | 0.0287 |
| Hair loss (no. of hairs lost/assessment) | 18.0 (6.5) | 14.8 (5.9) | 14.8 (4.6) | 16.8 (3.8) | 21.5 (5.4) | 0.0189 |

\* *p*-values calculated by χ2 test for gender and one-way ANOVA for baseline values.

### Sebum content

Group A (placebo) showed no consistent reduction in sebum production over the study period, with mean values fluctuating near baseline through D42 before declining slightly by D56 (Figure 2). In contrast, Groups B to E all exhibited progressive, significant reductions in sebum content beginning as early as D14 (C and D, p < 0.05; E, p < 0.01) with separation from placebo persisting and deepening through the remainder of the study period. By D56, Groups C, D, and E continued to exert significant reduction in sebum relative to placebo (C, p < 0.05; D, p < 0.01; E, p < 0.001). Within-group analysis confirmed that all four active treatment groups achieved statistically significant reductions in sebum relative to their own D14 baseline at later time points (Figure 2 bottom). For Groups B to E, significant within-group reductions were apparent from the D14-to-D28 interval onward, with all active groups reaching p < 0.0001 for the D14 vs. D56 comparison. Group A showed more modest but nonetheless statistically significant within-group changes at D28 vs. D42 and D42 vs. D56 (p < 0.05 and p < 0.01, respectively), consistent with a degree of natural sebum regulation over the study period.

**Figure 2.**
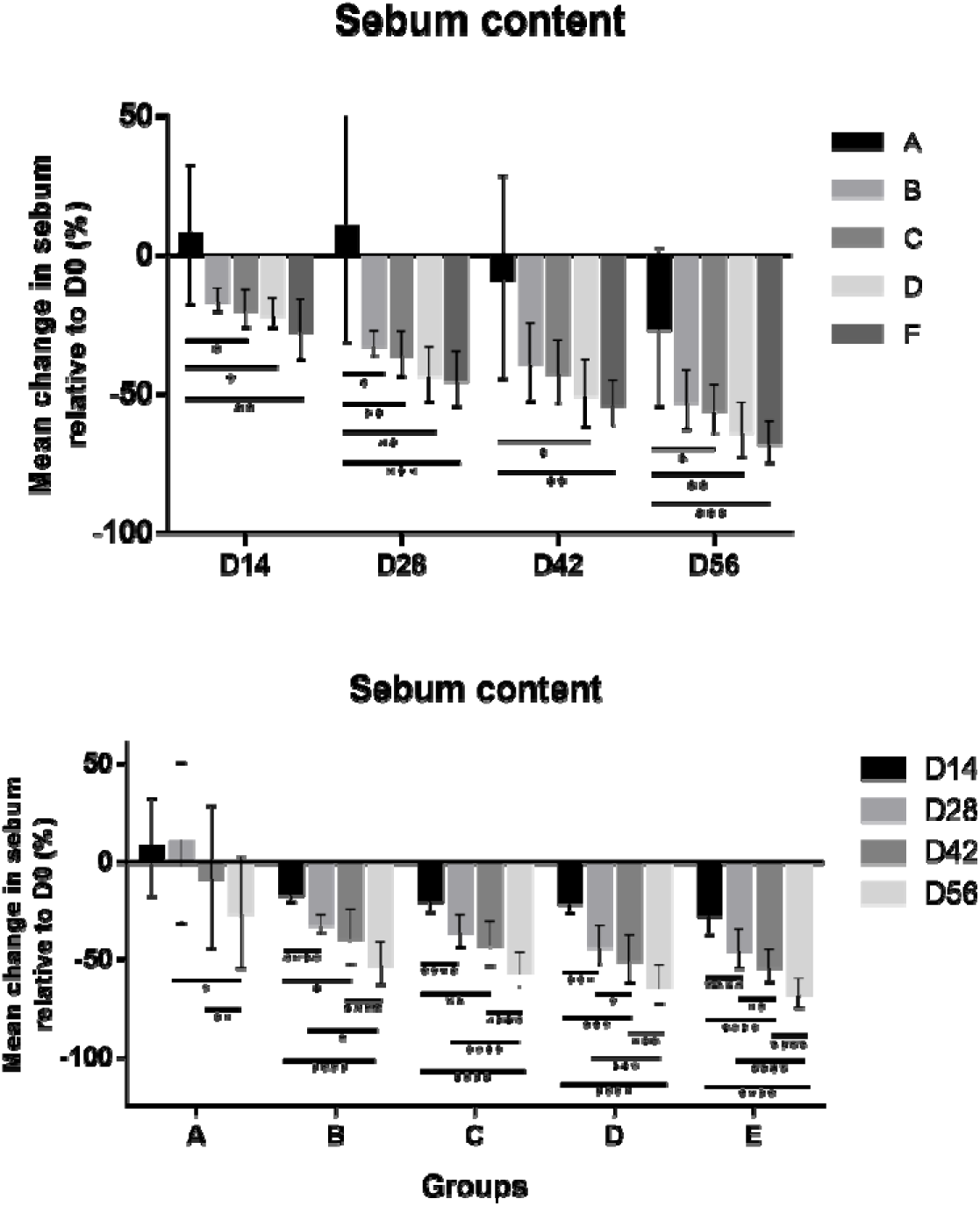
Change (%) in sebum content compared to baseline (D0) at D14, D28, D42, and D56. Top: Comparison between groups at the same timepoint; Bottom: Comparison between different timepoints of the same group. Comparison of sebum content at each timepoint between groups was performed using one-way ANOVA followed by Tukey’s HSD test; Comparison of within-group change over time was performed using repeated measures ANOVA followed by Tukey’s test. Results are presented in mean values with error bars representing 95% confidence intervals. * p < 0.05, ** p < 0.01, *** p < 0.001, **** p < 0.0001.

### Hair length

As expected for natural hair growth, all groups demonstrated increasing cumulative hair length over the study, but the magnitude of growth diverged substantially by treatment (Figure 3). By D14, Groups C to E all showed significantly greater hair length gain than placebo (C, p < 0.0001; D, p < 0.001; E, p < 0.0001), with Group E additionally exceeding Groups B and D at this early timepoint (B, p < 0.0001; E, p < 0.05). This separation widened over time: by D28, the mean cumulative increase in hair length for Group E (1.90 cm) was significantly greater than all other groups (all p < 0.0001). By D56, Group E showed the greatest mean cumulative hair length increase (3.86 cm), significantly outperforming all other groups (all p < 0.0001), while Group D (3.06 cm) was also significantly greater than Groups A (p < 0.05) and B (p < 0.01). As hair grows continuously, the within-group mean weekly change in hair length was used for comparison instead of the within-group mean cumulative hair length. Analysis of within-group mean weekly hair length change (Figure 3 bottom) revealed that all groups showed an increase in growth velocity over the study period, with Group E showing the most dramatic progression from 0.92 cm/week at D14 to 1.41 cm/week at D56 (p < 0.0001). Group A, while showing the smallest absolute velocity, nonetheless achieved statistically significant within-group increases in weekly growth rate between D14 and D42 and between D14 and D56 (both p < 0.01), possibly due to natural anagen phase turnover during the study period. Groups C, D, and E all showed extensive within-group significance across multiple timepoint comparisons (p < 0.01 to p < 0.0001).

**Figure 3.**
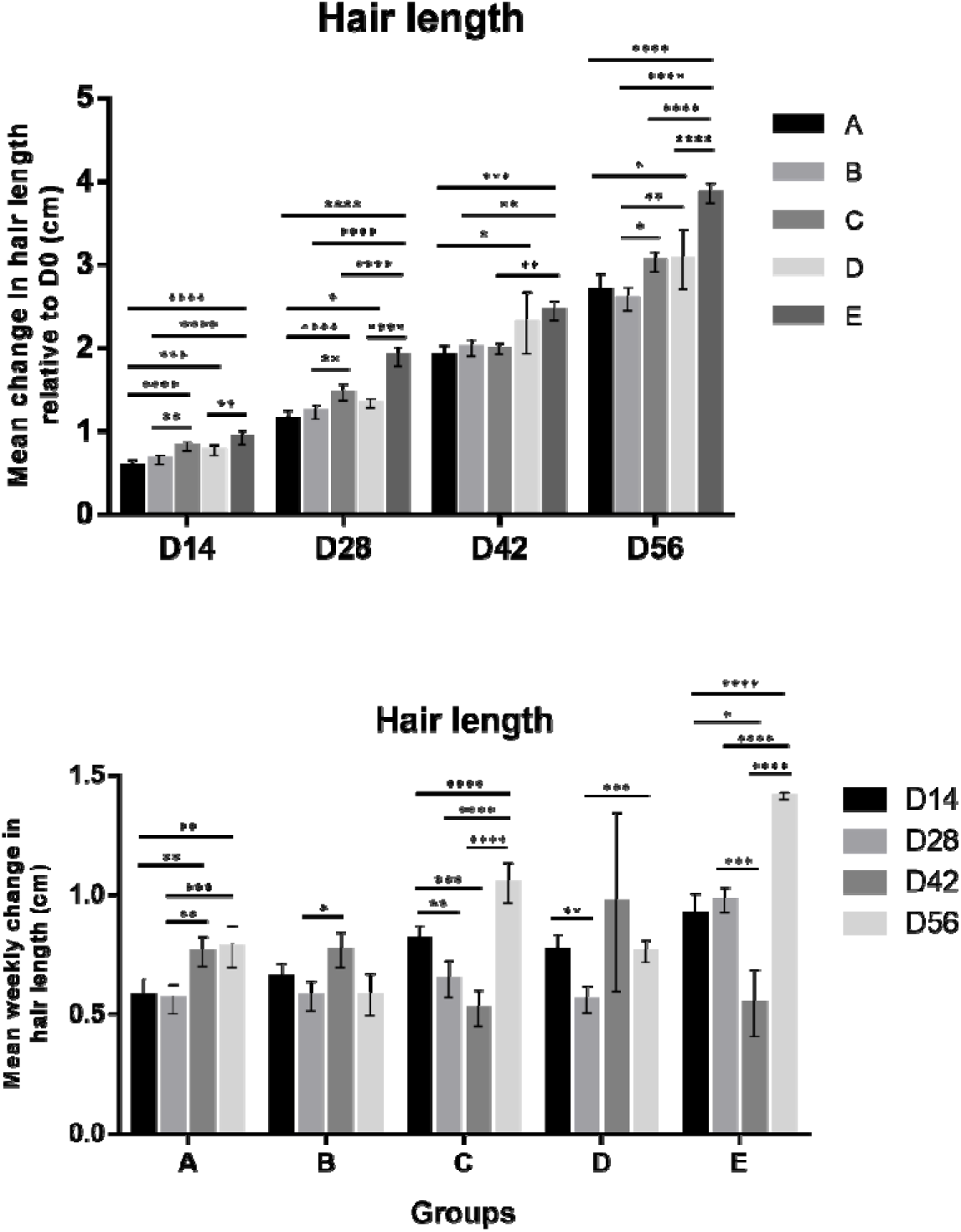
Change (cm) in hair length compared to baseline (D0) at D14, D28, D42, and D56. Top: Comparison between groups at the same timepoint; Bottom: Comparison of mean change in weekly hair growth between different timepoints of the same group by subtracting mean hair length between two timepoints. Comparison of hair length between groups at each timepoint was performed using one-way ANOVA followed by Tukey’s HSD test; Comparison of within-group weekly change in hair length over time was performed using repeated measures ANOVA followed by Tukey’s test. Results are presented in mean values with error bars representing 95% confidence intervals. * p < 0.05, ** p < 0.01, *** p < 0.001, **** p < 0.0001.

### Hair density

Group A showed no meaningful or significant change in hair density at any timepoint, with values remaining flat or slightly negative throughout the study (Figure 4). All active treatment groups showed positive increases from D14onward, with Groups C, D, and E differing significantly from Group A at every time point assessed (D14 through D56, p < 0.01 to p < 0.0001). Among the active groups, E achieved the largest density gains (up to 17% above baseline), significantly over Group B at D28 (p < 0.001) and D42 (p < 0.0001). In within-group analysis (Figure 4 bottom), significant progressive density increases were observed in Groups B (D42 vs. D56, p < 0.01), C (D14 vs. D28, p < 0.001; D28 vs. D42, p < 0.05), D (D14 vs. D28, p < 0.001), and E (D14 vs. D28, p < 0.0001; D14 vs D56, p < 0.05; D42 vs D56, p < 0.05). Again, Group A showed no significant within-group differences at any timepoint.

**Figure 4.**
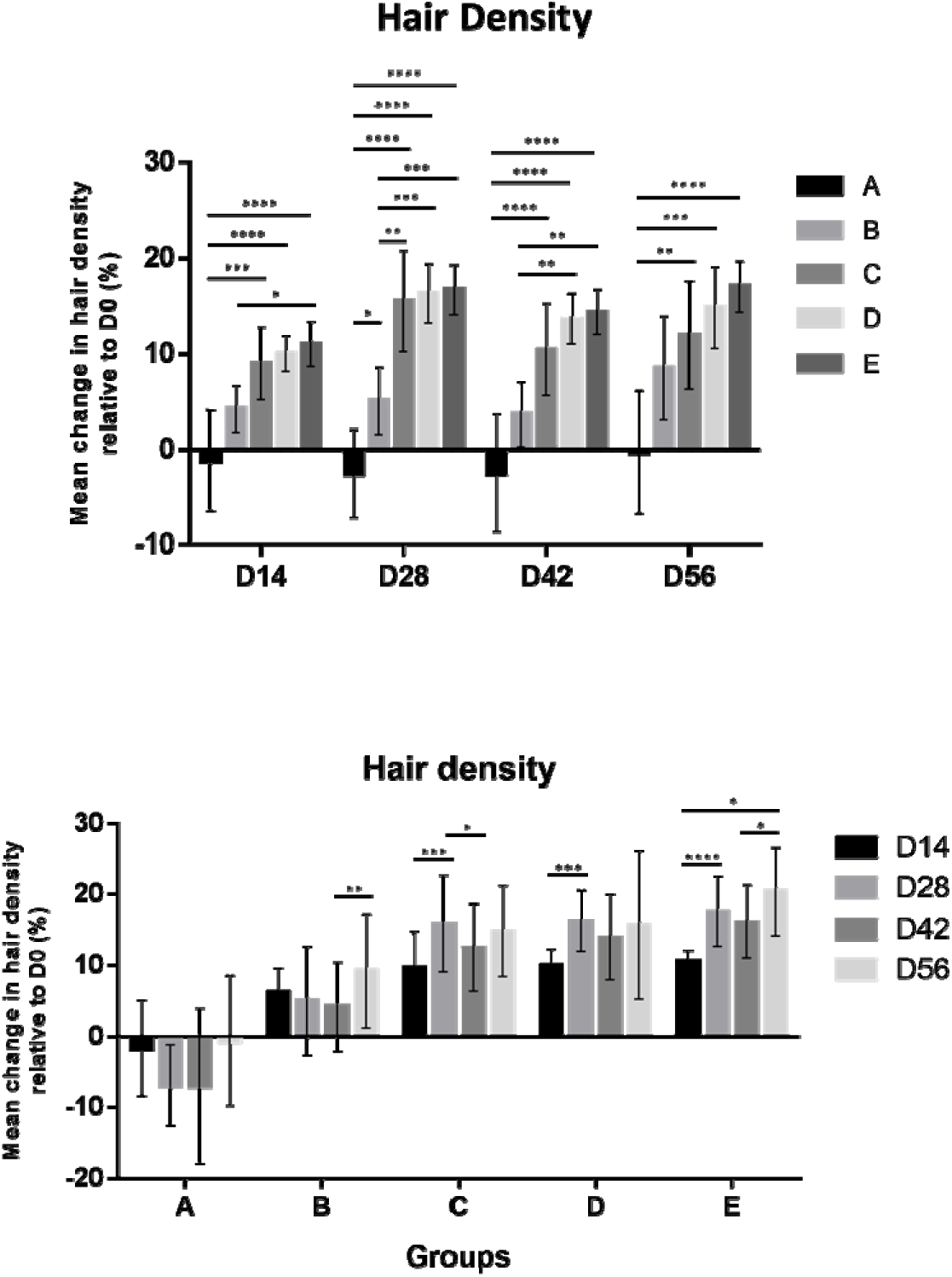
Change (%) in hair density compared to baseline (D0) at D14, D28, D42, and D56. Top: Comparison between groups at the same timepoint; Bottom: Comparison between different timepoints of the same group. Comparison of ha r density content at each timepoint between groups was performed using one-way ANOVA followed by Tukey’s HSD test; Comparison of within-group change over time was performed using repeated measures ANOVA followed by Tukey’s test. Results are presented in mean values with error bars representing 95% confidence intervals. * p < 0.05, ** p < 0.01, *** p < 0.001, **** p < 0.0001.

### Hair thickness

Hair thickness increased progressively across all active treatment groups while Group A remained virtually unchanged throughout the study (Figure 5). Significant differences from Group A were evident as early as D14 for all active groups (B, p < 0.05; C, p < 0.001; D and E, p < 0.0001) and persisted through D56. Group E resulted in the greatest increase in hair thickness at every timepoint, reaching a mean of 18.7 _μ_m above baseline by D56, significantly exceeding Groups A, B (both p < 0.0001), and C (p < 0.001). Within-group comparison (Figure 5 bottom) showed that Groups B, D, and E all achieved significant increases in hair thickness relative to their own D14 values across multiple timepoint comparisons. It is also notable that Groups C to E all achieved significant increases at D42 vs. D56 (D and E, p < 0.01; C, p < 0.0001).

**Figure 5.**
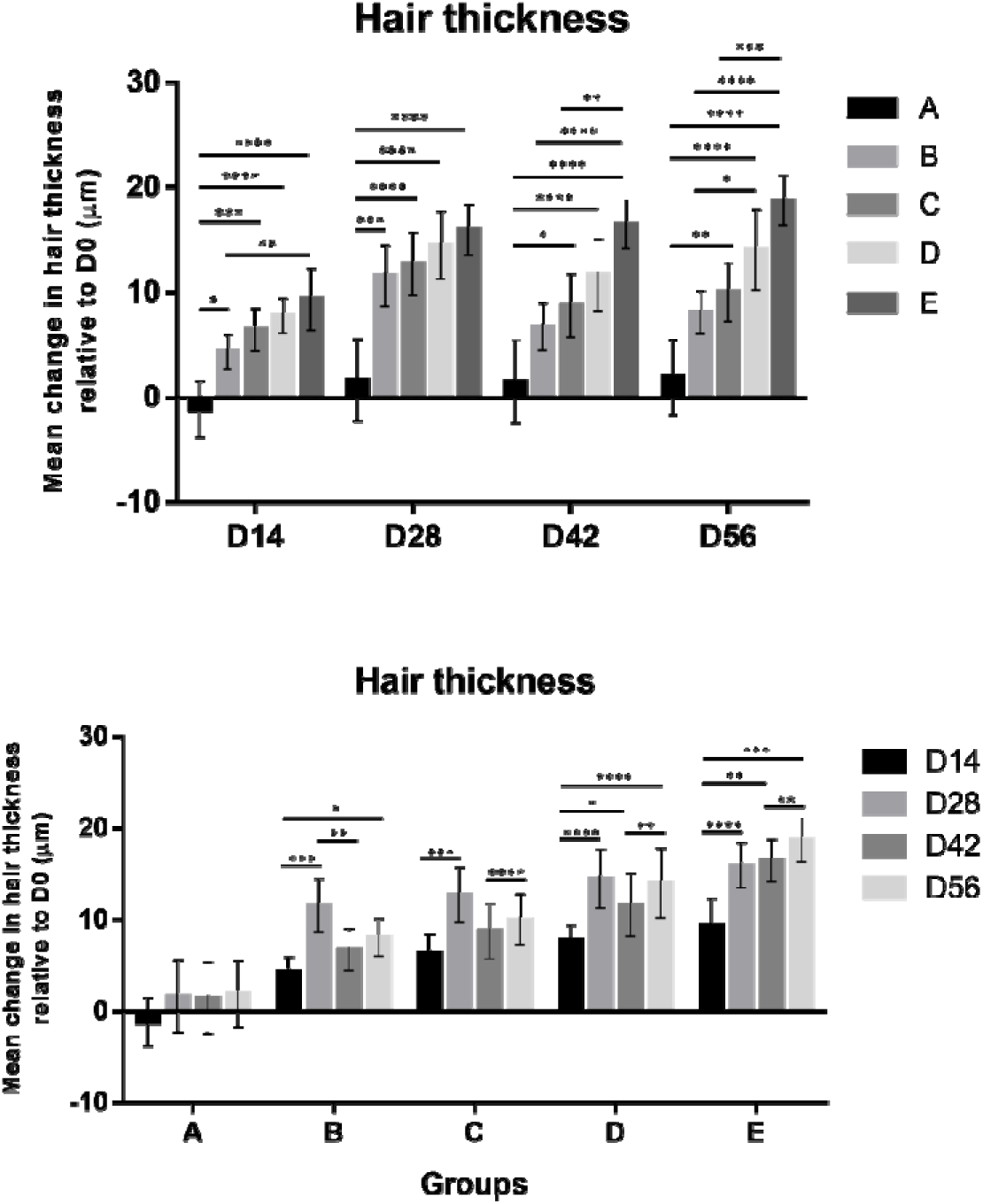
Change (µm) in hair thickness content compared to baseline (D0) at D14, D28, D42, and D56. Top: Comparison between groups at the same timepoint; Bottom: Comparison between different timepoints of the same group. Comparison between groups at each timepoint was performed using one-way ANOVA followed by Tukey’s HSD test; Comparison of within-group change over time was performed using repeated measures ANOVA followed by Tukey’s test. Results are presented in mean values with error bars representing 95% confidence intervals. * p < 0.05, ** p < 0.01, *** p < 0.001, **** p < 0.0001.

### Hair loss

Reduction in hair loss only became apparent progressively across the course (Figure 6). At D14, significant differences have yet to be observed, while at D28, Groups D and E showed significantly greater reductions in hair loss than Group A (both p < 0.05). By D42, this pattern broadened, with Groups C (p < 0.01), D (p < 0.01), and E (p < 0.001) all significantly outperforming Group A, and Group E additionally exceeding Group B (p < 0.05). By D56, the treatment effect was most pronounced: Group E cut hair shedding by almost half (approximately −48% from baseline), significantly outperforming Groups A (p < 0.0001), B (p < 0.0001), and C (p < 0.05). Within-group analysis (Figure 6 bottom) demonstrated that Groups B to E all achieved significant, progressive reductions in hair loss relative to their D14 values, with significance spanning multiple timepoint pairs from D28 onward (p < 0.05 to p < 0.0001 for D14 vs. D56 comparisons in all active groups). Placebo had no significant within-group change at any timepoint.

**Figure 6.**
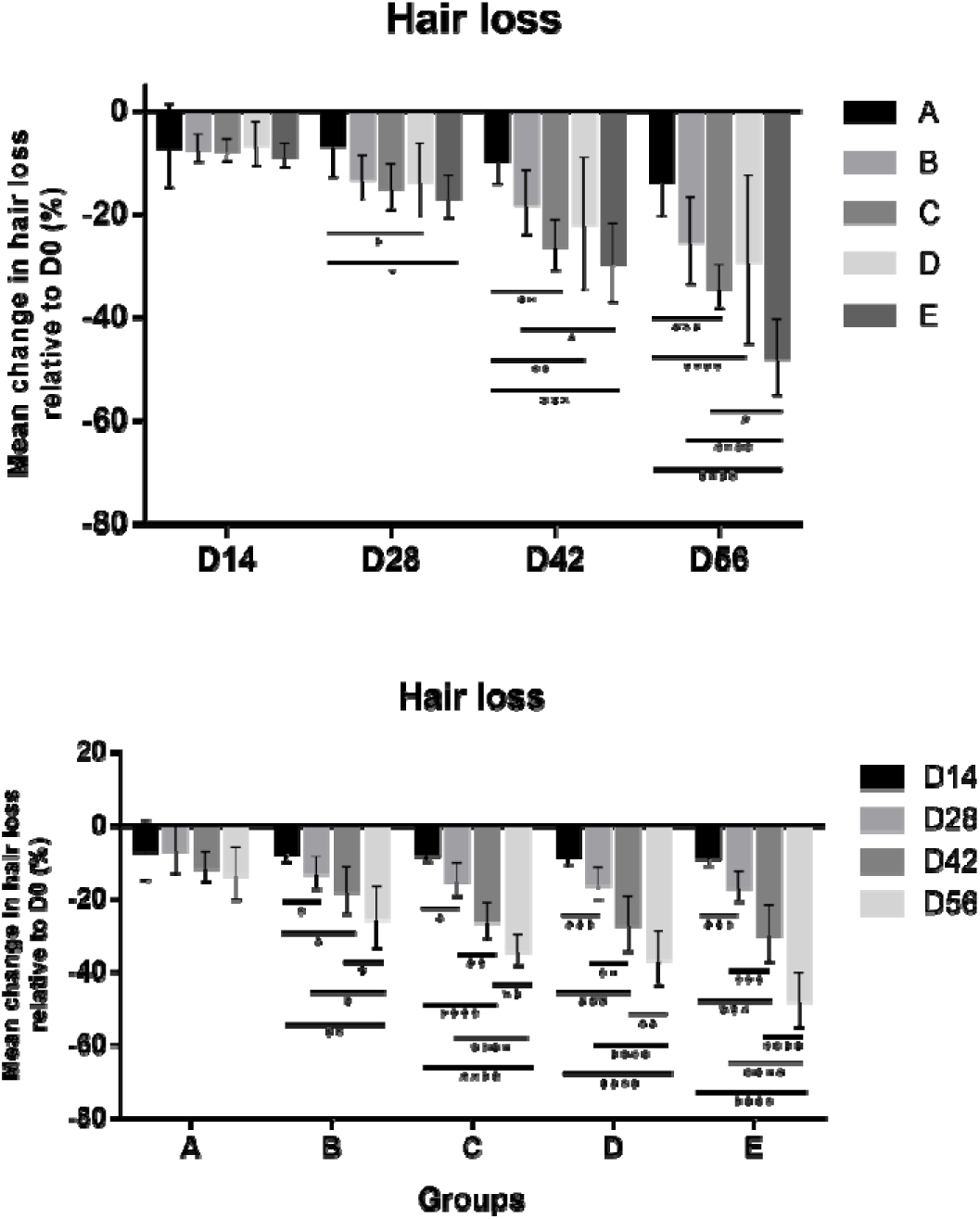
Change (%) in hair loss (shedding) compared to baseline (D0) at D14, D28, D42, and D56. Top: Comparison between groups at the same timepoint; Bottom: Comparison between different timepoints of the same group. Comparison of hair loss at each timepoint between groups was performed using one-way ANOVA followed by Tukey’s HSD test; Comparison of within-group change over time was performed using repeated measures ANOVA followed by Tukey’s test. Results are presented in mean values with error bars representing 95% confidence intervals. * p < 0.05, ** p < 0.01, *** p < 0.001, **** p < 0.0001.

Photographs from participants of Group E taken at D0 (pre-treatment) and D56 are shown in Figure 7, demonstrating visibly increased hair density, particularly at the vertex region.

**Figure 7.**
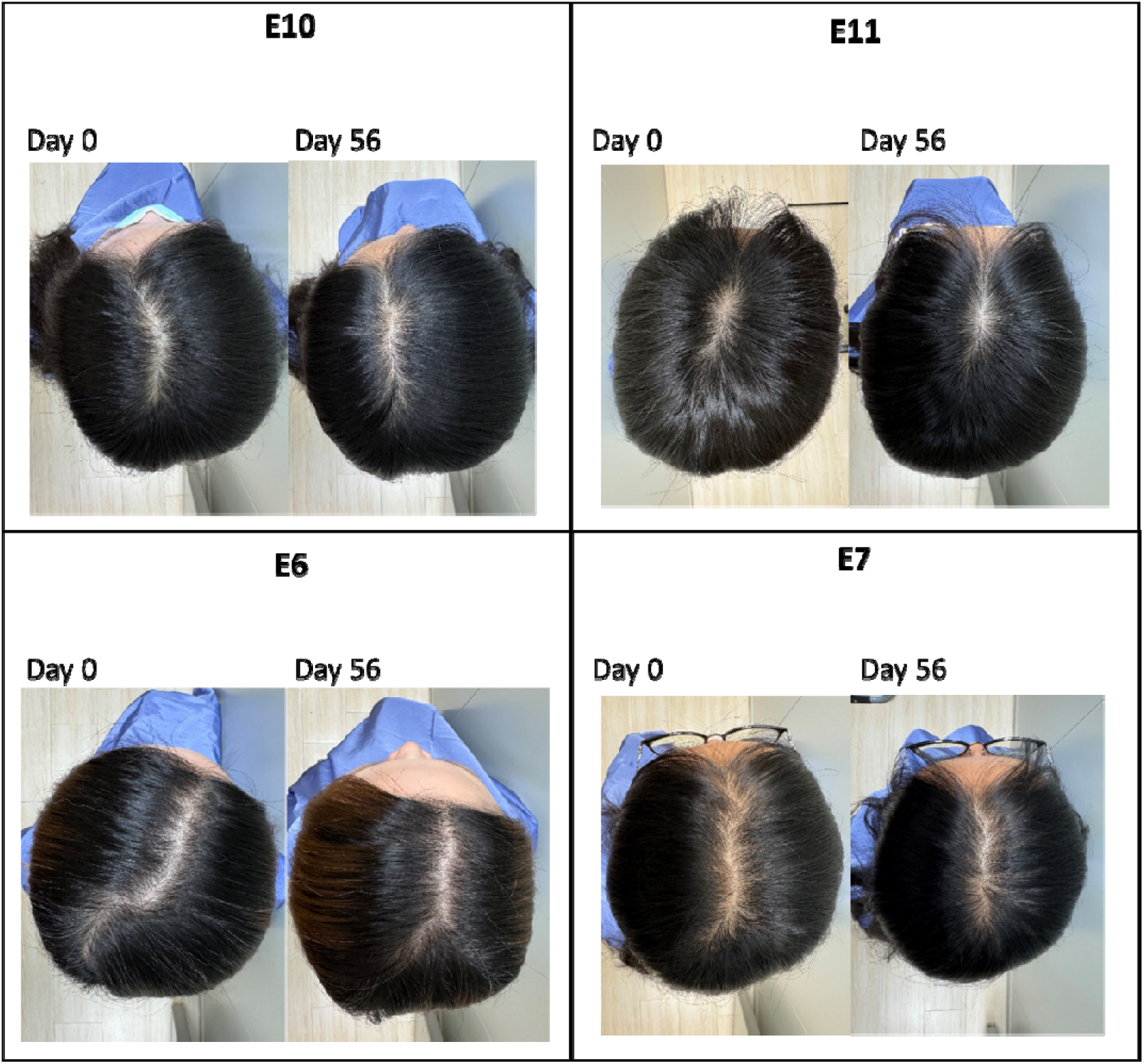
Representative images of the participants in Group E taken from above (vertex) on D0 and D56.

## Discussion

This 56-day randomized, placebo-controlled trial evaluated five topical formulations of increasing complexity across five scalp and hair endpoints. All active groups significantly outperformed placebo on most parameters from Day 28 onward, with effects progressively increasing through Day 56. Group E, the formulation combining *C. asiatica* EV, ginger-derived EV (GDEV), rIGF-1, and rFGF-7, produced the most consistent and comprehensive benefits across all endpoints. These findings both confirm and extend those observed in our previously published pilot study, and their comparison offers insight into the added value of the dual-EV architecture, the mechanistic synergism of the formulation’s components, and the role of seasonal follicular biology in shaping outcome magnitudes.

### Analysis of Results

Excessive sebum production contributes to scalp microbiome dysbiosis, follicular obstruction, and inflammation, which negatively impact hair growth[28]. In this study, Group E achieved a 67% sebum reduction from baseline by Day 56, versus 26% in the placebo, while Groups B to D also showed significant reductions from Day 28 onward. The mechanistic basis for sebum suppression in Group E likely involves synergistic pathways: *C. asiatica* EVs containing triterpenoids such as asiaticoside and madecassoside, which suppress inflammation via COX-2 and iNOS downregulation as previously observed *in vitro*; GDEVs containing NF-κB-suppressive miRNAs that downregulate IL-6, IL-8, and TNF-α[29]; the anti-inflammatory and antioxidant effects of IGF-1 via PI3K/AKT[30]; and caffeine’s suppression of excess sebum production by androgenic signaling[31,32]. The convergence of these distinct mechanisms in Group E explains why neither growth factors alone (Group C) nor EVs alone (Group D) achieved comparable sebum suppression.

All active groups demonstrated progressively greater cumulative hair length than the placebo group, with Group E reaching 3.86 cm by Day 56 and significantly outperforming all other groups. Hair growth velocity was analyzed using the weekly growth rate, which revealed that Group E showed a progressive increase in velocity through Day 56, while Groups C and D showed intermediate patterns and Group B tracked more closely to placebo. FGF-7, as a paracrine growth factor secreted by dermal papilla cells, activates keratinocyte proliferation (hence the alternate name keratinocyte growth factor, KGF) and anagen induction via MAPK/ERK, PI3K/AKT, and Wnt/β-catenin pathways. Both IGF-1 and FGF-7 converge on ERK and AKT signaling to promote epithelial regeneration and maintain the anagen phase, and IGF-1 prolongs anagen by suppressing catagen-associated apoptosis through Bcl-2 upregulation and boosting perifollicular angiogenesis via VEGF[11,14]. Panthenol is the precursor to vitamin B5 (pantothenic acid) and contributed incrementally by upregulating Ki-67, β-catenin, and VEGF in outer root sheath cells while suppressing TGF-β1-mediated apoptotic signaling[20].

Hair density and/or thickness followed similar trends. Group E produced the largest gains in both parameters, significantly outperforming placebo at all time points from Day 14 onward, whereas Groups C and D achieved intermediate effects. Placebo remained essentially unchanged on both measures throughout the study. The progressive increase in hair thickness across all active arms confirms that even the base formulation with caffeine and panthenol provides support to the follicle. Only Group E, which also contained twice the concentration of growth factors used in Group C, showed a statistically significant within-group change in hair density from Day 14 to Day 56. These results may also reflect VEGF upregulation enhancing perifollicular angiogenesis by *C. asiatica*[33] and activation of crucial anagen and follicular stem cell cycling signaling[34].

Hair loss reduction emerged later than structural improvements, with significant differences from the placebo first appearing at Day 28. By Day 56, Group E reduced hair shedding by almost half and significantly outperformed all other groups. The latency of this effect relative to length and thickness could be explained by the fact that hair shedding occurs over multiple follicular cycles, as compared to the more immediate effect on hair structure of actively growing follicles in anagen.

### Dual Plant-EV Formulation and the Contribution of Ginger-Derived EVs

The most important difference from the prior published trial is the incorporation of GDEVs alongside *C. asiatica* EVs in Groups D and E. To our knowledge, this is the first clinical trial to evaluate a dual plant-derived EV formulation containing growth factors in a topical application for scalp and hair health, with two distinctive EV profiles to achieve broader coverage of the pathways implicated in scalp and follicular health.

GDEVs have been identified as containing approximately 125 distinct miRNAs, high concentrations of 6-gingerol and 6-shogaol, and immunomodulatory lipids involved in counteracting inflammation by downregulating NF-κB, IL-6, IL-8, and TNF-α[29]. Ginger constituents also induce Nrf2 nuclear translocation and activate Wnt/TCF4 signaling in stem cells, important in this context as bulge-resident hair follicle stem cells are responsible for follicular renewal[35]. Given Wnt/β-catenin signaling’s central role in hair follicle stem cell activation and anagen induction, the capacity of GDEVs to engage this pathway independently of the growth factor components provides a plausible mechanism for their inclusion[34]. A recent review by Hao et al[15]. proposed GDEVs as a natural therapeutic candidate for alopecia; thus, our strategy of including GDEV in formulation is well-founded[15]. Interestingly, 6-gingerol by itself has an inhibitory effect on hair growth, as demonstrated by Miao et al[36]., who showed that free 6-gingerol suppresses hair shaft elongation and dermal papilla cell proliferation *in vitro* and *in vivo* via a pro-apoptotic decrease in the Bcl-2/Bax ratio. However, GDEVs are fundamentally distinct from plant extracts such as free 6-gingerol, as phytochemicals are delivered along with miRNAs and anti-inflammatory lipids as a heterogeneous signaling complex, thus making the pathways and mechanisms complicated and not attributable to any single component. Bioavailability, intracellular concentration, and receptor-level context differ substantially between EV-encapsulated and free-compound delivery, and the superior clinical data from Group E over other formulations provided empirical evidence that the GDEV-containing formulation promotes rather than suppresses hair growth under the conditions studied.

The mechanistic complementarity of the two EV species with growth factors was the probable basis for the superior performance of Group E over both Group C (growth factors only) and Group D (EVs only). However, because Group E contained twice the concentration of rIGF-1 and rFGF-7 compared with Group C while the EV concentration was unchanged relative to Group D, the superior performance of Group E cannot be attributed solely to the combination of growth factors and EVs and may partly reflect the higher growth factor dose. *C. asiatica* EVs target TGF-β/Smad-mediated collagen synthesis, COX-2-mediated inflammation, and sebaceous gland regulation via triterpenoid cargo and regulatory mRNAs[18,37]. GDEVs target NF-κB-mediated cytokine production, Nrf2-mediated antioxidant defense, and Wnt-driven stem cell activation. While IGF-1 and FGF-7 trigger signaling pathways such as MAPK/ERK, PI3K/Akt, and Wnt/β-catenin to promote stem cell activation, inhibit apoptosis, prolong follicular growth, and initiate transition into anagen[11,14]. All of these individual components engage and modulate a complex network of pathways to provide a favorable follicular microenvironment.

### Comparison with the Prior Trial and the Role of Seasonal Hair Cycling

Although this was not designed as a confirmatory study, the current results are consistent with the prior publication, reinforcing the robustness of the multi-component formulation[19]. Granted that the two studies could not be directly compared, the absolute magnitudes of several endpoints differ in ways not attributable to formulation alone, and we propose that the different enrollment seasons of the two trials may have played a role in the differences between the two studies. Although human scalp hair cycling is largely asynchronous, annual periodicity of growth has been documented, as in the landmark study by Randall and Ebling[38]. The study demonstrated that in the Northern Hemisphere, the anagen proportion peaks above 90% in March and declines to a nadir in September, with daily shedding approximately doubling from winter to the August-to-September peak. Another trichographic analysis of 823 women confirmed maximal telogen rates in summer and a secondary, less pronounced peak in spring, with the lowest telogen rates in late winter[39]. The changes are proposed to result from photoperiod-mediated changes in hormone levels, with rising day length in spring that activates prolactin-mediated reactivation of telogen follicles and anagen induction, while the declining period of daylight in autumn favors catagen entry and telogen accumulation[40]. Sebum production is also well-documented to be lower in winter and higher in summer, driven by temperature, humidity, and UV-mediated stimulation of sebocyte activity[41,42]. The prior study began shortly after the anagen peak in April and progressed into July, the period of naturally rising telogen rates and shedding, which was identified as the anagen nadir in the Randall and Ebling study. The present trial began at the post-autumn shedding peak in November and traversed the full winter anagen recovery and spring anagen maximum before concluding in May. These nearly opposite seasonal trajectories create systematically different interpretive contexts for each outcome. However, as the trials were conducted in a subtropical setting in Taiwan, the magnitude of seasonal follicular variation was likely more subtle relative to the temperate Northern European populations in which the previous large population datasets were generated, though measurable seasonal hair cycling in East Asian subtropical cohorts has been documented which also showed increase rate of telogen around August to September but to a lower extent[43].

For hair length, the current study’s slightly larger gains in Group E (∼3.9 vs. ∼3.5 cm) are consistent with amplification by the spring anagen peak, during which the highest proportion of follicles is actively producing hair shaft. For hair density, the current study showed modestly attenuated Group E gains (∼17% vs. ∼23.9%). When all groups (including placebo) benefit from the natural winter-to-spring increase in the anagen proportion, the contribution of active treatment is harder to resolve against the growing background. In the April to July study, active groups maintained or improved density against a declining seasonal background, producing a larger and more clearly treatment-attributable separation. The most pronounced discrepancy in hair loss between the two studies (∼48% vs. ∼64% reduction in Group E), a seasonal explanation is plausible: In the April to July study, participants’ shedding was naturally progressing toward its summer peak, meaning the placebo group’s shedding either worsened or remained elevated, making active groups’ reductions attributable to treatment. In the present study, shedding naturally declined from the autumn peak to lowest in winter, thus raising the background and reducing the apparent effect of the active formulation’s contribution. Crucially, the fact that Group E consistently outperformed all other groups across all parameters in both trials further strengthens the argument for formulation efficacy rather than mere seasonal artifact.

### Comparison with Current Clinical Literature in Exosomes for Alopecia

A recent systematic review by Al Ameer et al[44].synthesized 11 clinical studies, including two randomized clinical trials (RCTs), which investigated MSC-derived exosome interventions across 298 patients with diagnosed AGA and other forms of alopecia. Across the included studies, hair density gains ranged from 9.5 to 35 hairs/cm² and hair thickness increases reached up to 13 µm, with the largest gains observed in the highest-quality RCT using adipose-derived MSC exosomes delivered via intradermal microneedling. The approximately 19 µm hair thickness gain achieved by Group E in the present study and the 27.9 µm reported in our prior published trial[19] have shown non-inferiority and are even superior to the results with MSC exosomes, despite being achieved through once-daily non-invasive topical self-application. The present formulation is further distinguished by its plant-derived EV source. In the systematic review, all but one study used human or animal MSC-derived exosomes, and only one pilot RCT evaluated any plant-derived exosome preparation, albeit a single-species botanical extract rather than isolated, characterized plant EVs[44]. Therefore, the current trial is among the few placebo-controlled RCTs utilizing rigorously defined plant-derived EVs, and the first to combine two distinct plant EV species with recombinant growth factors in a topical formulation. The review also noted that its included studies enrolled patients with established, diagnosed alopecia, whereas the present trial targeted healthy adults without clinically diagnosed hair loss, reflecting a preventive and cosmetic maintenance use.

### Limitations

Several limitations should be acknowledged here. As in the previous study, the participants were predominantly female (∼83% per group) and relatively young, limiting generalizability to older male populations with age-related alopecia. The sample size was relatively small, with 12 participants per group; this remained an exploratory study, which precludes subgroup analyses by sex, age, or baseline scalp condition. The 56-day duration, while sufficient to detect meaningful differences, is short relative to the full human anagen phase (2–6 years) and does not address durability of effects after discontinuation, which is a recognized concern for pharmaceutical treatments such as minoxidil[45]. Additionally, despite blok randomization, several baseline values differ significantly as in Table 2: hair thickness (*p*=0.0158), hair density (*p*=0.0287), and hair loss (*p*=0.0189), which reflect the inherent challenge to achieve a completely equal baseline with small group sample size of *n*=12. To minimize this effect, we normalized the outcome measurements to each participant’s own baseline, although we acknowledge that these pre-existing differences may nonetheless have influenced the magnitude of changes observed in the study. As an example, Group E started the study with the highest mean hair loss count (21.5 hairs/assessment), which was higher than Groups B and C (both 14.8) while Group E’s superior reduction in hair loss remains internally consistent and statistically robust. On the other hand, Groups D and E began with lower mean hair thickness than Groups A and B, and yet produced the greatest absolute thickness gains, suggesting that the observed treatment effects represent an authentic response rather than an artifact resulting from baseline. Importantly, Group E contained twice the concentration of rIGF-1 and rFGF-7 compared with Group C, with the EV concentration unchanged relative to Group D. The superior effects of Group E therefore reflect the combined influence of the EVs and a higher growth factor dose, and this design cannot separate the contribution of growth factor dose from that of the growth factor-EV combination. Finally, the absence of mechanistic biomarker or histological data (scalp biopsy, follicular transcriptomics, serum IGF-1, or microbiome profiling) prevents direct confirmation of the proposed signaling mechanisms, although the above would require invasive procedures.

### Conclusions

Here we demonstrated that the dual plant-EV combination of *C. asiatica* EV and GDEV with rIGF-1, rFGF-7, caffeine, and panthenol produced the most comprehensive and progressive improvements in all parameters evaluated compared to other formulations. To our knowledge, this is the first clinical trial to deploy two distinct plant-derived EV populations in combination with recombinant growth factors for topical hair and scalp care. The consistent directional replication of key findings from the prior published trial, demonstrated across opposite seasonal backgrounds, strengthens confidence in the biological activity of the treatment effect and supports the continued development of this naturally derived, non-pharmaceutical formulation class as a scalp and hair health intervention.

## Author Contributions

Conceptualization, supervision and project administration: T.-M.C., C.C., T.-Y.K. Visualization and formal analysis: T.-M.C., W.-J.H., L.T.-C.L. Investigation, data acquisition and curation: T.-M.C., H.-C.H., J.-Y.L. Methodology: T.-M.C., C.-C.W., H.-C.H., J.-Y.L., Y.-S.C., P.-L.K., W.-H.T., W.-Y.Q., I.P., T.-Y.K. Resources: C.-C.W., Y.-S.C., P.-L.K., W.-H.T., T.-Y.K. Writing—original draft preparation: T.-M.C., W.-J.H., L.T.-C.L. Writing—review and ed-iting: T.-M.C., W.-J.H., L.T.-C.L., W.-Y.Q., I.P., C.C., T.-Y.K. All authors reviewed the manuscript.

## Institutional Review Board Statement

The finalized protocol, case report form, advertisement, and informed consent form were reviewed and approved by the institutional review board of the Antai Tian-Sheng Memorial Hospital (IRB No. 25-090-A, approved 21 October, 2025). All participants provided written informed consent prior to the start of the study. This study was conducted in compliance with the Declaration of Helsinki and in compliance with all International Conference on Harmonization Good Clinical Practice Guidelines. This clinical study was registered on ClinicalTrials.gov under NCT07271212 on 09 December, 2025.

## Data Availability Statement

All relevant data is provided within the manuscript.

## Acknowledgments

We thank colleagues at Schweitzer Biotech Company for re-viewing and providing feedback for the manuscript preparation.

